# Performing Qualitative Mask Fit Testing without a Commercial Kit: Fit Testing Which can be Performed at Home and at Work

**DOI:** 10.1101/2020.08.04.20168344

**Authors:** Eugenia O’Kelly, Anmol Arora, Charlotte Pearson, James Ward, P. John Clarkson

## Abstract

**Introduction:** Qualitative fit testing is a popular method of ensuring the fit of sealing face masks such as N95 and FFP3 masks. Increased demand due to the COVID-19 pandemic has led to shortages in testing equipment and has forced many institutions to abandon fit testing. Three key materials are required for qualitative fit testing: the test solution, nebulizer, and testing hood. Accessible alternatives to the testing solution have been studied. This exploratory qualitative study evaluates alternatives to the nebulizer and hoods for performing qualitative fit testing.

**Methods:** Four devices were trialed to replace the test kit nebulizer. Two enclosures were tested for their ability to replace the test hood. Three researchers evaluated promising replacements under multiple mask fit conditions to assess functionality and accuracy.

**Results:** The aroma diffuser and smaller enclosures allowed participants to perform qualitative fit tests quickly and with high accuracy.

**Discussion & Conclusion:** Aroma diffusers show significant promise in their ability to allow individuals to quickly, easily, and inexpensively perform qualitative fit testing. Our findings indicate that aroma diffusers and homemade testing hoods may allow for qualitative fit testing when conventional apparatus is unavailable. Additional research is needed to evaluate the safety and reliability of these devices.

## Introduction

Qualitative fit testing provides the ability to ensure acceptable fit of a sealing face mask such as an N95 or FFP3. Unfortunately, during the COVID-19 pandemic, a dramatic increase in the demand and use of protective equipment and fit testing equipment, coupled with severe supply shortages, has forced many institutions to abandon fit testing ^1^.

Fit is a primary factor in determining whether a mask is capable of reducing the spread and inhalation of fine particles. Previous studies have noted that even if the materials of a mask have high filtration efficiency, the effectiveness of the mask is hampered by an imperfect seal. A study by Huff et al, using nebulized radioactive technetium, found that an ineffective seal is the principal cause of airborne contamination among those wearing respirators ^2^. This finding is supported by the findings of Cooper et al, who found that leakage around masks accounted for one-third of the airflow across the mask for surgical masks and one-sixth of the flow for respirators ^3^.

Qualitative testing is most commonly adopted to assess the fit of half-face respirators such as N95 and FPP3 masks ^4^ as it is less expensive, more accessible, faster, and less demanding of staff time than the alternative of quantitative fit testing ^4^ Qualitative testing is widely used in hospitals, where a high throughput rate is critical for facility performance ^5^.

Under normal circumstances, the equipment required for qualitative fit testing is affordable and accessible. The tests usually require three items: a wearable testing hood; a testing solution aerosoliser; and an aerolised test solution. Unfortunately, disruption to supply chains and a surge in demand has limited their availability. Those seeking to purchase such equipment currently face out of stock notices or wait times, in the US, of up to and over eight weeks.

Solving the fit testing supply crisis is critical in order to enable hospitals and businesses to properly protect their workers. Prior studies have already shown the feasibility of making a homemade fit testing solution ^6,7^. This initial exploratory study aims to assess the effectiveness of alternatives to the mechanical elements of the qualitative fit test: the fit testing hood and nebulizer.

## Methods

### Testing and Verification

Three testers underwent quantitative fit tests to assess the fit of the masks on their face before assessing qualitative fit methods. Quantitative fit testing, which measures the number of particles inside and outside of a face mask, is a highly accurate means of measuring fit and is more accurate than qualitative fit testing ^6,8^. Results from the quantitative fit tests were used to determine what sensation (taste vs no taste) would be considered the correct response in the qualitative fit testing. Quantitative fit tests were conducted with a Portacount 8038+ using OSHA protocol 29CFR1910.134.

One member of the team performed initial testing. Additional testers were brought in to confirm and assess promising devices. More information can be found in the online data supplement.

### Testing Solution

Fakherpour et al and Mitchell et al have developed effective sodium saccharine testing formulas which can easily be made at home ^6,9^ Our formula was based on these studies. The testing solution consists of 830mg sodium saccharine to 100mL distilled water. Our sensitivity/threshold solution contained a sodium saccharine concentration between Fakherpour et all’s and Mitchel et all’s threshold solution, using 415mg sodium saccharine per 100ml. A nebulizer was used to ensure each test subject could detect the saccharine solution.

### Devices

Four devices were tested for their ability to replace the bulb nebulizer used for qualitative test kits. These included an ultrasonic mist maker (from AGPTEK, headquarters Guangdong, China), a spray bottle, a miniature wand humidifier (from JISULIFE, headquarters Guangdong, China), and an essential oil diffuser (from VicTsing, headquarters Sunnyvale, California USA). Flow rate information was not available for the spray bottle or miniature humidifier. The mini humidifier by JISULIFE packaging stated a diffuse rate of 25-40mL/h. The ultrasonic mist maker by AGPTEK claims to produce at least 400mL/h.

Devices were run for 60 seconds, or in the case of the spray bottle, for up to 10 sprays. If no taste could be detected within that period, the test condition was judged as ‘no taste’.

### Testing Enclosures

In qualitative testing, a hood with a small circular opening for ventilation and insertion of the squeeze nebulizer is used to concentrate the testing mist. Like the rest of the testing equipment, these hoods are expensive and, under these pandemic conditions, difficult to obtain. We tested two alternative testing enclosures to replace the hood: a clear storage cube measuring 11.81” square and a sturdy 2-gallon plastic bag. When testing the hood, which is designed for a horizontally discharging aerosolizing device, we used a piece of curved PVC fit to the hole in the testing hood and help direct the mist from the vertically discharging devices.

### Mask

Two different masks were worn during testing, an N95 mask manufactured by 3M and a KN95 mask from a Chinese manufacturer.

To assess the ability of the device and enclosure to accurately predict fit issues, five tests were conducted in different fit states. Tests were performed with the unmodified N95 and KN95. The wearer then performed three additional tests with the N95 while intentionally causing a fit gap. For one test they placed the tip of their finger between the mask and the skin beneath their eye, causing a gap in the nose area. For the other test, they placed the tip of their finger between the bottom of the mask and their chin area, causing a chin gap. Finally, they placed the tip of their finger between the cheeks and edge of the mask. These three tests enabled us to assess how well the testing setup could detect specific fit issues.

## Results

### Aerosolization Devices

The testers were able to taste the sensitivity solution with three of the devices used (see figure 1). The wand humidifier proved difficult to use and did not produce a clear taste sensation. The mist maker and spray bottle produced a taste sensation but had undesirable side effects such as causing the mask to become wet. The aroma diffuser produced accurate responses. The accuracy using this aroma diffuser was in line with the reported accuracy of commercial qualitative fit testing kits^6^.

**Figure 1:**
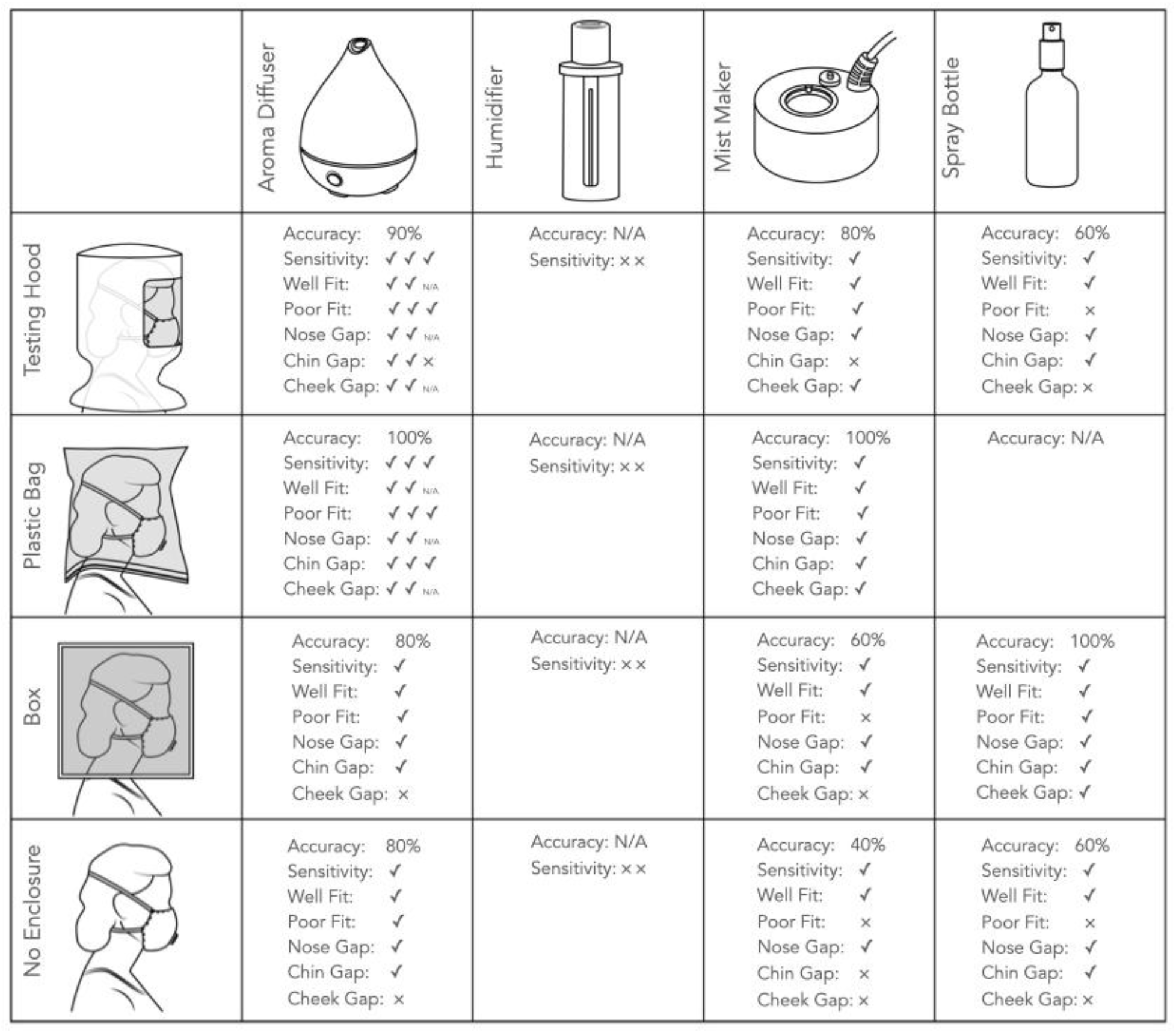
A chart of the fit test results with various diffusers and enclosures. A checkmark indicates the correct response to the testing condition while an x indicates an incorrect response to the test condition. Each column of marks indicates a different researcher’s responses.

### Testing Enclosures

Smaller testing enclosures allowed for faster detection of the testing solution in some, but not all, devices. The spray bottle did not function well in the hood and was unable to fit into the 2-gallon bag.

The smaller testing enclosures benefited both the accuracy and the detection speed when using the aroma diffuser and ultrasonic mist maker. The smallest enclosure, the plastic bag, produced the fewest inaccurate results and allowed for correct detection in the shortest period of time. Larger testing enclosures incurred greater inaccuracy and longer times to detection.

## Discussion

### Summary of Key Findings

Our results indicate that qualitative fit testing can be conducted effectively using inexpensive homemade testing solutions and household testing devices. Moreover, a simple, quick, and easily replicated at-home or work setup may enable users to test for proper fit of sealing face masks (see figure 2). We found that aroma diffusers and zipper storage bags were the most accurate alternatives to typical solution nebulisers and testing hoods respectively. The degree of success, reliability and ease of set up for our homemade testing apparatus exceeded our expectations and suggests that alternative testing enclosures and nebulisers may prove useful where conventional apparatus is unavailable or unaffordable. However, it must be emphasized that this research is preliminary. The method proposed is not NIOSH compliant. We cannot be sure if this method of qualitative testing is as effective or safe as standard NIOSH testing methods using commercially available equipment. Repurposing equipment for qualitative fit testing may come with risks to the wearer. We advise anyone who intends to use this method to proceed with caution and at their own risk.

**Figure 2:**
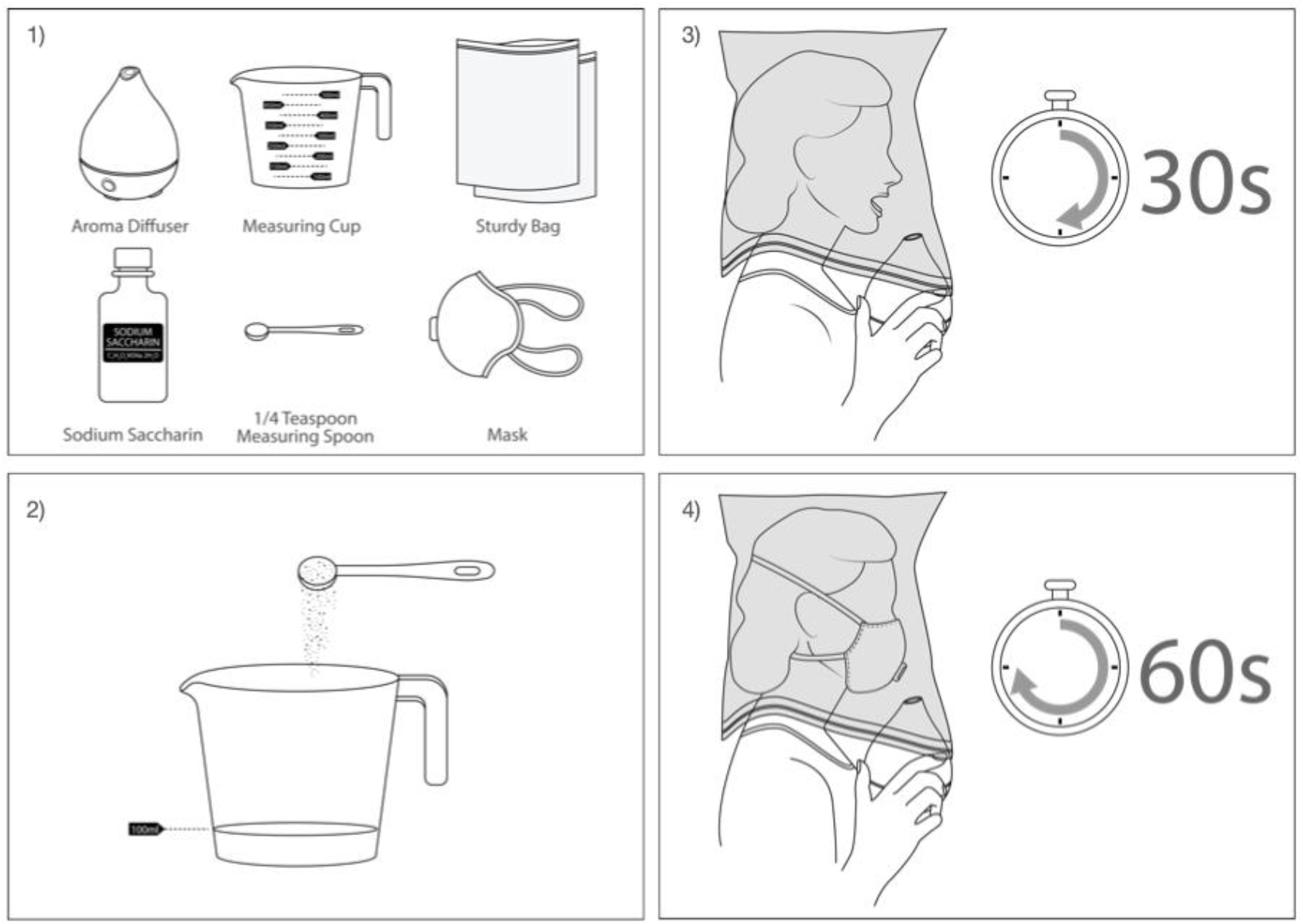
The procedure followed to assess qualitative fit testing mechanisms, shown with the highly successful aroma diffuser and sturdy bag. In step 1, the essential equipment was gathered. The required 830mg of sodium saccharin proved to fill slightly less than ¼ a teaspoon. In step 2, 100ml of water was mixed with ¼ of sodium saccharin. Sensitivity or threshold solution contained half the amount of sodium saccharine. In step 3, the participant donned the enclosure and breathed as instructed in OSHA qualitative fit testing instructions, with the mouth partly open and tongue placed towards the front of the mouth. If the sensitivity solution could not be tasted in 30 seconds, the sensitivity test was failed, and the test discontinued. In step 4, the participant donned the mask and performed each fit test procedure for 60 seconds, taking breaks in between tests for additional safety.

### Poorly Functioning Devices & Enclosures

We do not recommend the use of the spray bottle or the ultrasonic mist maker for fit testing as both made the mask visibly wet and consequently compromised the future use of the mask.

Our results indicate that humidifiers may not be fit for quantitative testing. While it is possible the wand humidifier we tested provided an insufficient flow rate and that a more powerful humidifier might be adequate, it is likely that filters included with most humidifiers removed enough of the sodium saccharin to prevent the device from working as a fit testing device. The aroma diffuser was effective at testing masks without an enclosure but improved with a smaller space. Passing the aroma diffuser around the face so that the fog comes in contact with the mask improves the accuracy when no enclosure is used. The box we used provided little benefit over no enclosure. It was not small enough nor airtight enough to create a concentration of aerosolized sodium saccharine around the mask area. A smaller or more airtight box might be more effective.

### Successful Devices & Enclosures

Commercial aroma diffusers proved highly effective in aerosolizing the testing solution.

Small enclosures, or those most similar to the conventional testing hood, produced the fastest and most accurate results when using an aroma diffuser.

If a testing hood is not available, we recommend using a robust freezer zipper storage bag with at least a two-gallon capacity. A large freezer plastic bag, kept open and placed over the head, proved an inexpensive and highly effective option. Due to the possible risk of suffocation, it is vital that extra caution is taken while using a plastic bag in this manner. This type of enclosure should only be used by risk-aware adults who are able to quickly and safely maneuver the bag on and off. Only plastic bags with heavier structure, such as Ziplock freezer bags (from SC Johnson, headquarters in Racine, Wisconsin, USA), should be used. Their more rigid form and heavy plastic zipper resists deforming and blocking the air flow, nose, or mouth. Bags should never be sealed around the neck. The entirety of the bottom of the bag should be left open to promote airflow. The weight and structure also help prevent the bag from becoming tangled with the testing equipment or closing around the tester’s neck. Light, poorly structured bags, such as plastic takeout or grocery bags, garbage bags, or product packaging bags should never, under any circumstances, be used as these bags have a higher chance of causing suffocation. Bags should always be large enough to provide at least 2 inches in front of the mask when worn. For safety, a second adult should be present and aware at all times the bag is worn and able to quickly remove the bag in case of any issue. Testing with any enclosure not approved for qualitative fit testing may be hazardous and is undertaken at the risk of the user.

## Conclusion

Despite being less accurate than quantitative fit testing, qualitative testing remains a vital tool to ensure mask-wearers are protected by assessing mask fit. It is currently more important than ever that people have access to inexpensive fit-testing protocols so that they can protect themselves as well as others.

Our initial tests indicate that replacing the expensive test kit nebulizer with an aroma diffuser may be feasible. The accuracy of our tests were in line with, and often exceeded, the accuracy of qualitative fit tests performed with commercial solutions and equipment ^6^. This new qualitative testing method shows promise but requires additional research to ensure its effectiveness and safety. Given the growing risk the SARS-CoV-2 poses around the world, some may judge the potential risk of using this unproven testing method less severe than the risk of wearing a respirator whose poor fit does not protect the wearer. Future work is needed to assess the safety and efficacy of using alternative methods, as well as their ability to function with other testing solutions including bitter solutions and alterative testing solutions such as aloe vera^10^.

## Data Availability

Data will be available as an online supplement or in an open data repository.

## Acknowledgements

We would like to give special thanks to Dr Ming Jeffrey Kao, who introduced us to the problem which motivated this study. We would also like to thank Corinne O’Kelly for making this research possible.

